# Registry Forge: an open-source end-to-end pipeline for patient-directed SMART on FHIR registries

**DOI:** 10.64898/2026.06.02.26354637

**Authors:** Danielle Boyce, Alan Premasiri, Shawn Sullivan, Beth Levine, Fernando G. Vieira

## Abstract

**Objectives:** Patient-directed SMART on FHIR lets registries acquire longitudinal electronic health record data, but the payload requires substantial engineering before use. We present Registry Forge, an open-source pipeline that converts it into research-ready outputs.

**Materials and Methods:** Registry Forge decodes and parses mixed C-CDA, HTML, RTF, PDF, and FHIR inputs, joins records to a canonical patient identifier, and emits a browser-viewable dashboard, an OMOP CDM v5.4 data set, GA4GH Phenopackets v2, a code inventory, and regex extractions of disease-specific narrative content.

**Results:** Applied to the ALS Research Collaborative Study (94 participants, 56 US health systems), it processed 22,686 source files and 1,791 FHIR Bundles (109,599 resources); only 15.0% of files were full C-CDA.

**Discussion:** This pipeline generalizes to any registry acquiring data through patient-directed SMART on FHIR.

**Conclusion:** Registry Forge closes the acquisition-to-analysis gap with no server infrastructure and is openly available.

**LAY SUMMARY:** Patient registries and natural history studies (referred to as ‘registries’ in this article) are research databases that collect data on people with a particular medical condition, often over time. To work well, they need detailed medical records, either from the participant answering surveys or through information from each participant’s doctors. A federal rule lets participants log in to their hospital’s patient portal and grant a registry access to their own electronic health record. This solves the data collection problem but creates a new one: the downloaded files arrive in many formats and have to be cleaned, combined, and standardized before researchers can use them. Small registries, especially those for rare diseases, often cannot afford the engineers required to do this work.

We built Registry Forge, a free open-source software tool that does this work automatically. We used it for the ALS Research Collaborative Study, a long-running natural history study of amyotrophic lateral sclerosis run by the ALS Therapy Development Institute. Registry Forge processed records from 94 study participants whose care spanned more than 50 US health systems and turned them into research-ready data sets. The tool runs on a standard laptop and is free for anyone to use.

## BACKGROUND AND SIGNIFICANCE

The Substitutable Medical Applications, Reusable Technologies (SMART) on FHIR specification describes how a third-party application authenticates to a patient’s electronic health record (EHR) account and retrieves clinical data on the patient’s behalf using OAuth 2.0.[1,2] Following the 21st Century Cures Act Final Rule, certified health IT modules that meet the ONC standardized API criterion must support FHIR R4 APIs and SMART App Launch and OpenID Connect authorization for patient-facing access.[3,4] A patient can therefore authorize an external registry to retrieve available clinical data exposed through a compliant patient-facing API, reducing the need for separate site-by-site data use agreements.

Acquiring data is not the same as having usable data. In our patient-directed retrieval workflow, FHIR Bundle exports and DocumentReference and Binary retrieval yielded a mixture of Consolidated Clinical Document Architecture (C-CDA) XML, FHIR JSON, and narrative attachments in vendor-dependent formats.[5,6,7] Each artifact must be decoded, parsed, deduplicated, joined to a canonical patient identifier, and rendered for review. Small registries need outputs that registry leaders, clinical coordinators, curators, and investigators can read directly without running code, which argues for a deliberately wide, extract-all-curate-later pipeline coupled to a viewer the whole team can use.

## OBJECTIVES

To produce a reproducible, local-first, open-source pipeline that (i) decodes and parses every artifact in a SMART on FHIR-acquired clinical document repository, (ii) joins records across C-CDA and FHIR via a canonical patient identifier, (iii) enriches incomplete display-name fields, (iv) supports auditable exclusion of test records, (v) produces an OMOP Common Data Model (CDM) v5.4 data set tagged by Athena vocabulary release, (vi) produces GA4GH Phenopackets v2 documents, and (vii) recovers disease-specific content from unstructured note narratives.

## MATERIALS AND METHODS

### Inputs and architecture

Two parallel directories on local disk were processed: 22,686 XML files retrieved via SMART on FHIR DocumentReference and Binary endpoints, and 1,791 JSON files each containing a FHIR Bundle for one patient. Registry Forge is organized as a seven-stage core that emits a single JSON bundle, plus independent add-on modules that consume that bundle (Figure 1). The separation means an adopting registry that only needs a viewer runs only the core; adopters mix and match add-ons by use case. The pipeline follows an extract-all, curate-later philosophy: the same observation may appear in both C-CDA and FHIR streams, and rather than deduplicate aggressively at ingest, the bundle preserves both rows with source-system provenance tags so adopters apply their own rules downstream.

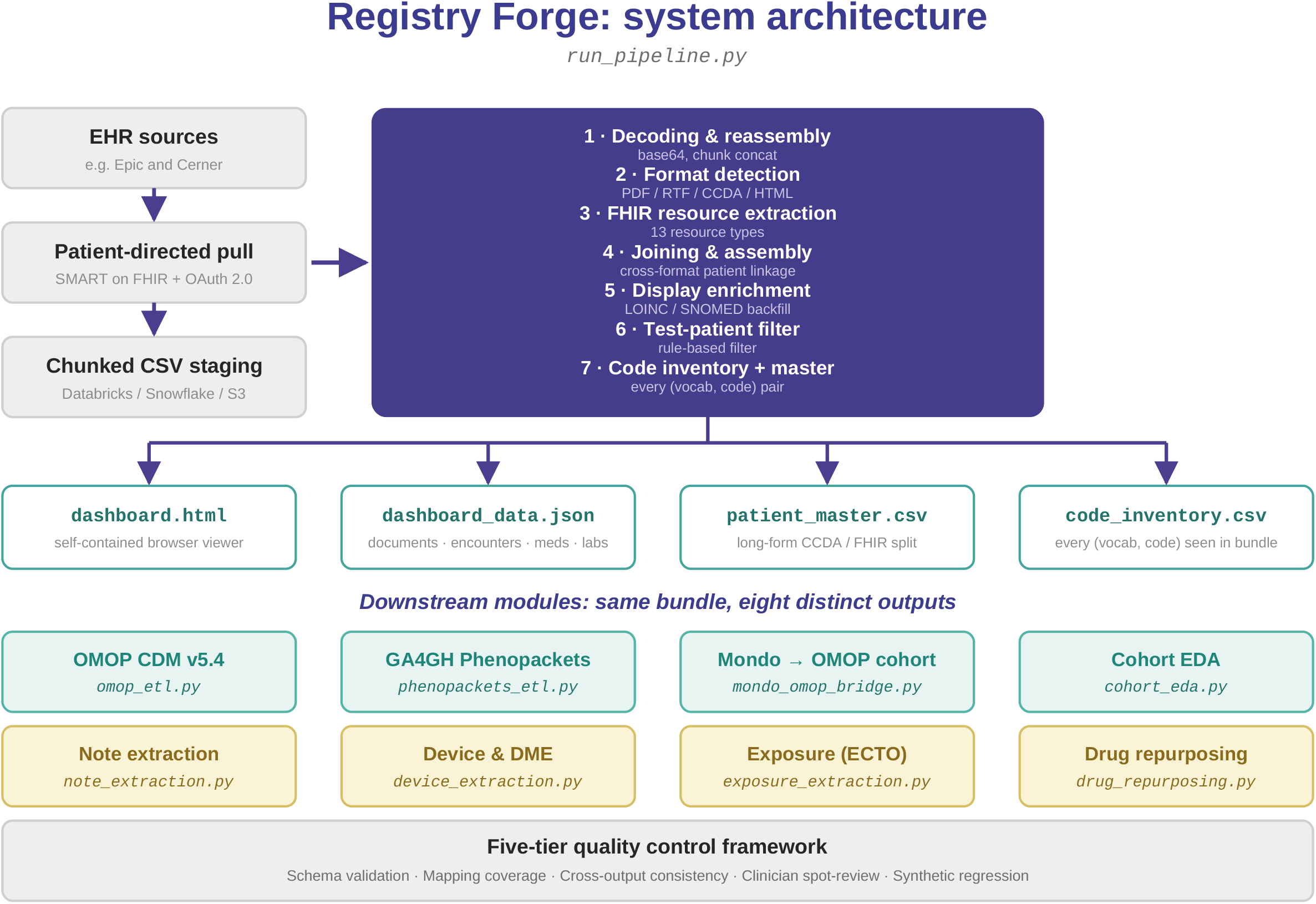

### Core pipeline

Stage 1 reassembles chunked CSV exports by document identifier and base64-decodes payloads with up to two passes. Stage 2 routes each file to a format-specific parser based on magic-byte detection: xml.etree.ElementTree for C-CDA, a custom stripper for RTF, pypdf for PDFs, and html.unescape with tag removal for HTML. A per-domain code prioritization (RxNorm > SNOMED > NDC for medications; SNOMED > ICD-10-CM for problems; LOINC > SNOMED for labs) selects the primary code while the complete coding list is preserved. Stage 3 walks each FHIR Bundle and dispatches each resource to a type-specific extractor across 13 resource types, resolving patient linkage via subject.reference. Stage 4 unifies structured tables by patient identifier, tags each row source=ccda or source=fhir, and emits the JSON bundle consumed by a static HTML dashboard (Figure 2). Stage 5 backfills missing displayName attributes from a built-in lookup of common LOINC codes. Stage 6 excludes test patient records via a configuration-driven filter supporting four match types. Stage 7 emits a code inventory listing each unique (vocabulary, code) pair with reference counts and patient counts.

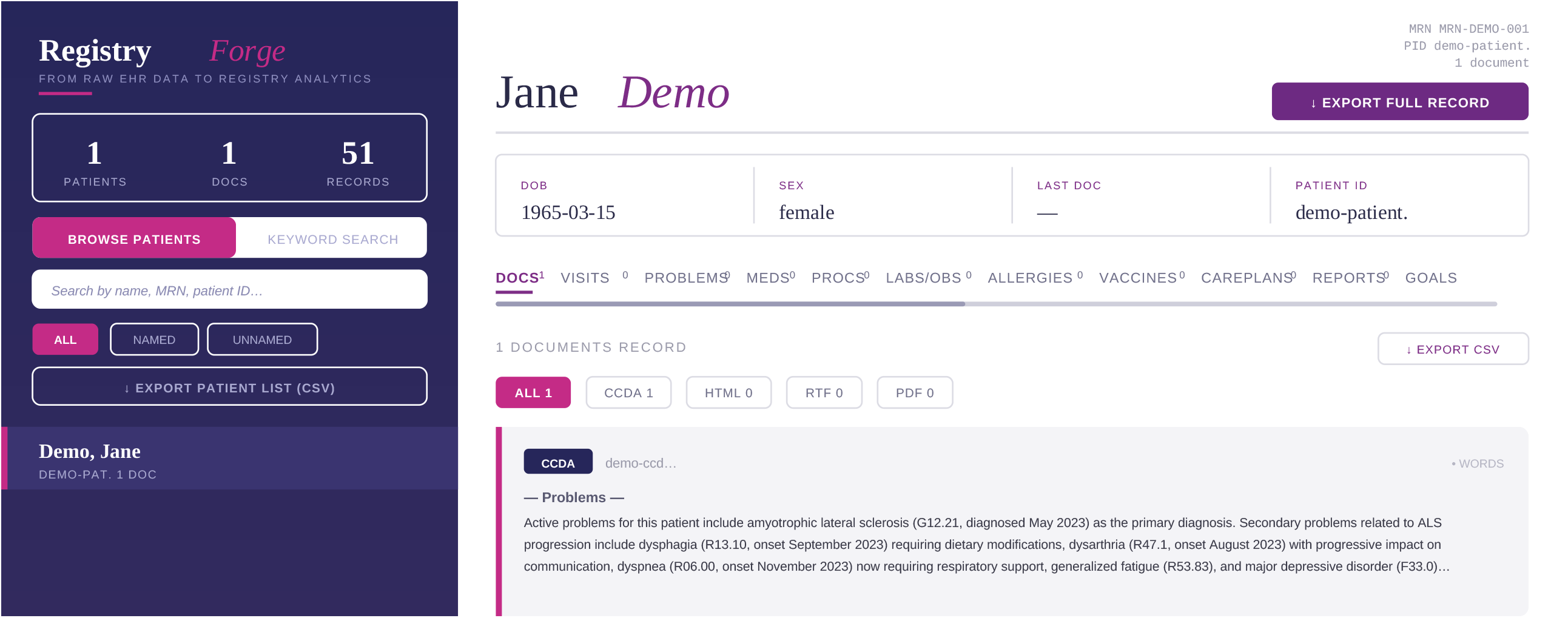

### Add-on modules

An OMOP ETL reads the bundle plus an OHDSI Athena vocabulary download and writes nine CDM v5.4 tables (PERSON, OBSERVATION_PERIOD, VISIT_OCCURRENCE, CONDITION_OCCURRENCE, DRUG_EXPOSURE, PROCEDURE_OCCURRENCE, MEASUREMENT, OBSERVATION, CDM_SOURCE), routing records by the domain_id of the standard concept rather than by source category.[8,9] A Phenopackets v2 ETL produces one JSON document per patient plus a Cohort document, with seed mappings for motor neuron disease, epilepsy, and autoimmune neurologic disease.[10] A regex note-extraction module applies a pattern library to every free-text source and emits one row per match with patient identifier, source, pattern, value, a snippet, and offset. Shipped ALS-specific patterns target ALSFRS-R scores, El Escorial certainty, site of onset, family history, gene mentions, forced vital capacity, and treatment milestones. The patterns are seed patterns for site-specific tuning, not validated production NLP; sites with mature NLP should use cTAKES,[11] medspaCy[12], or transformer-based models. Additional modules are documented in the public repository.

## RESULTS

We applied the pipeline to the ALS Research Collaborative Study EHR cohort (n=94 participants).[13] We attributed records to source systems using the FHIR service endpoint and resource metadata (Bundle.meta.source and entry.fullUrl), resolving named institutions where possible. Records resolved to 56 US health systems, predominantly Epic with some Cerner (Table 1). Generic Epic-hosted and Cerner tenant endpoints that could not be mapped to a specific institution were retained as unresolved source-system links. About one third of participants linked records from more than one system. Participants arriving without a name or source system in structured fields are flagged for review in the viewer; in our experience these details are often recoverable from the unstructured notes and can be corrected after import. Magic-byte format detection at Stage 2 revealed that only 15.0% of the 22,686 assembled files were full C-CDA XML documents; HTML narrative fragments (48.4%) and RTF clinical notes (34.3%) together comprised more than four-fifths of the repository, with PDF documents (2.3%) making up the remainder. A registry that assumes all DocumentReference attachments are C-CDA would silently drop the majority of its source material.

**Table 1.**
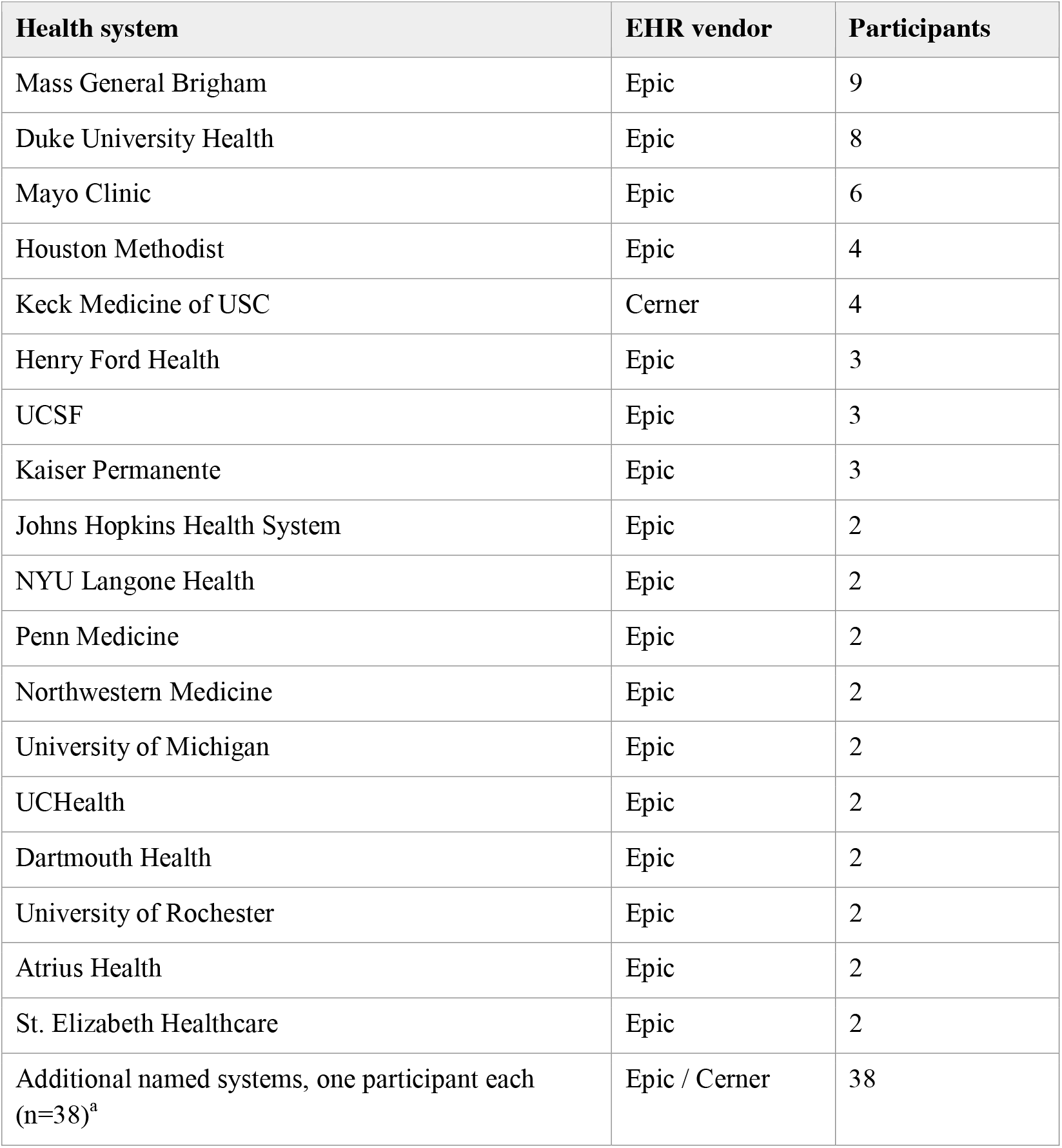
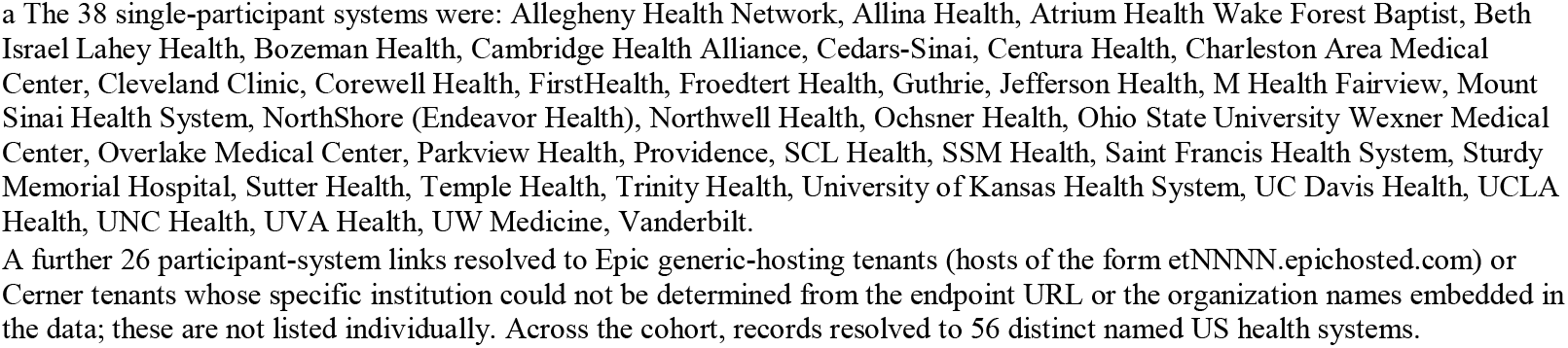
Health systems represented in the ALS Research Collaborative Study cohort, attributed from the FHIR service endpoint. Counts are distinct participants per system and are non-exclusive; some participants linked records from more than one system, so the column sums to more than the 94 distinct participants.

Across all sources, the pipeline processed 109,599 FHIR resources from the Bundle stream and extracted 33,084 C-CDA section narratives plus 14,825 narrative documents after test-record exclusion (791 records associated with two confirmed test patients were removed). The Stage 7 code inventory enumerated several thousand unique (vocabulary, code) pairs spanning SNOMED CT, ICD-10-CM, RxNorm, LOINC, CVX, and CPT-4. Approximately 36% of LOINC-coded observations in source C-CDA documents carried no displayName attribute. Because CDA displayName values are human-readable aids rather than the semantic source of truth, Registry Forge backfills labels for review while retaining the original code and provenance; this complements broader work on LOINC code and unit harmonization in real-world data.[14] Stage 5 enrichment populated 39,022 labels, raising display-name coverage from 64% to 84%. The OMOP ETL successfully populated all nine target CDM tables, and CDM_SOURCE.csv recorded the exact vocabulary versions used. The Phenopackets ETL produced one valid v2 JSON document per participant. On the synthetic demonstration cohort, the regex-based note-extraction module recovered every category of ALS-specific content patterned in the shipped library.

A fully synthetic single-patient interactive demonstration of the dashboard, exercising every record category with codes verified against RxNav, SNOMED CT, ICD-10-CM, LOINC, CVX, and CPT-4, is publicly hosted alongside the source code at https://alstdi.github.io/RegistryForgeALS/ to allow other registries to evaluate the pipeline before adopting it.

## DISCUSSION

The challenges of extracting and processing patient-directed EHR data are not unique to ALS or the ALS Research Collaborative Study. Any registry acquiring data through patient-directed SMART on FHIR will face structurally similar problems, and the broader literature on secondary use of EHR data has documented them across disease areas.[15,16] Table 2 summarizes the universal patterns and the corresponding pipeline layer that addresses each. What needs to be customized per registry is well-defined: a thin acquisition adapter to populate the two input directories; patient-identifier mapping queries against the registry’s own warehouse; disease-specific display-name lookups and regex patterns; and seed phenotype/disease tables for adopters using the Phenopackets module.

**Table 2.** Universal post-acquisition problems facing patient-directed SMART on FHIR registries and the Registry Forge layer that addresses each. Adapter effort indicates the work an adopting registry can expect to do for that pipeline layer beyond plugging the pipeline into the registry’s own acquisition output.

| Universal problem | Registry Forge layer | Adapter effort |
| --- | --- | --- |
| Heterogeneous payload formats (HTML / RTF / PDF behind DocumentReference) | Stage 2 magic-byte detection plus format-specific parsers | None; works as shipped |
| Identifier-mapping provenance gaps | Stage 4 join against patient-import audit table | Adopter writes SQL against own warehouse |
| Vendor-specific display-name omissions | Stage 5 LOINC / SNOMED display-name backfill | Extend lookup table for disease area |
| Test or non-clinical records | Stage 6 configuration-driven filter (four match types) | None; works as shipped |
| Source codes unknown to mapping pipeline | Stage 7 code inventory CSV | None; works as shipped |
| OMOP vocabulary version drift across runs | Vocabulary-tagged output folder plus CDM_SOURCE.csv versions | None; works as shipped |
| Disease-specific content trapped in narrative | Note-extraction module (regex library) | Author site-specific patterns |

Registry Forge has been developed and tested against data from a single registry and validated end-to-end on a fully synthetic demonstration cohort; validation against data sets from other registries and EHR vendors is invited. The production cohort reported here is small and reflects the early adoption phase of the ALS Research Collaborative Study’s SMART on FHIR onboarding; the patterns described should be read as engineering generalizations rather than as epidemiologic claims about ALS. The shipped regex pattern library is a seed for site-specific tuning, not validated production NLP. The five-tier quality-control framework included with the pipeline (schema validation, mapping-coverage tracking, cross-output consistency checks, periodic clinician spot-review, and synthetic-cohort regression testing) is a starting set and is not a substitute for the Kahn et al. data-quality framework[17] or the OHDSI Data Quality Dashboard.[18] Registry Forge automates the engineering work of registry data integration but does not replace the clinical, methodological, and editorial judgment adopting organizations must apply to their own data.

## CONCLUSION

Patient-directed SMART on FHIR has substantially expanded the data acquisition options available to patient registries, but the analytics work required to make acquired data usable falls disproportionately on small registries lacking in-house data engineering capacity. Registry Forge closes this gap end-to-end with no server infrastructure, converting a heterogeneous SMART on FHIR-acquired document repository into a researcher-usable browser viewer, an OMOP CDM v5.4 data set, GA4GH Phenopackets, a code inventory, and structured extractions of disease-specific narrative content. The full implementation, including a fully synthetic demonstration data set, is publicly available at https://alstdi.github.io/RegistryForgeALS.

## ACKNOWLEDGMENTS

The authors gratefully acknowledge the ALS Research Collaborative Study participants, without whom this study would not have been possible, and Unite Genomics.

## ETHICS

The ALS Research Collaborative Study operates under continuous Institutional Review Board approval from Advarra (IORG0000635) and is conducted under a single, ongoing protocol. The secondary data analysis activities described in this manuscript are within the approved protocol. No individual-level data are reproduced in this article; all figures depicting patient-level information are drawn from a fully synthetic demonstration cohort that contains no protected health information.

## DATA AND CODE AVAILABILITY

Source code for the complete pipeline, the browser dashboard, and a fully synthetic single-patient demonstration cohort are publicly available at https://alstdi.github.io/RegistryForgeALS/ (repository: https://github.com/alstdi/RegistryForgeALS) under the MIT License. The exact release used for this manuscript is archived with a citable DOI (https://doi.org/10.71944/2P5C-NG50) through ALS TDI’s DataCite membership. The clinical data described in this manuscript cannot be openly shared due to the terms of the ALS Research Collaborative Study informed consent. A de-identified version of the ALS Research Collaborative Study data are available to qualified researchers through the ARC Data Commons (https://www.als.net/arc/data-commons/).

## USE OF ARTIFICIAL INTELLIGENCE

A generative AI tool (Claude Opus 4.7) was used for copyediting and for generation of Figure 1.

---

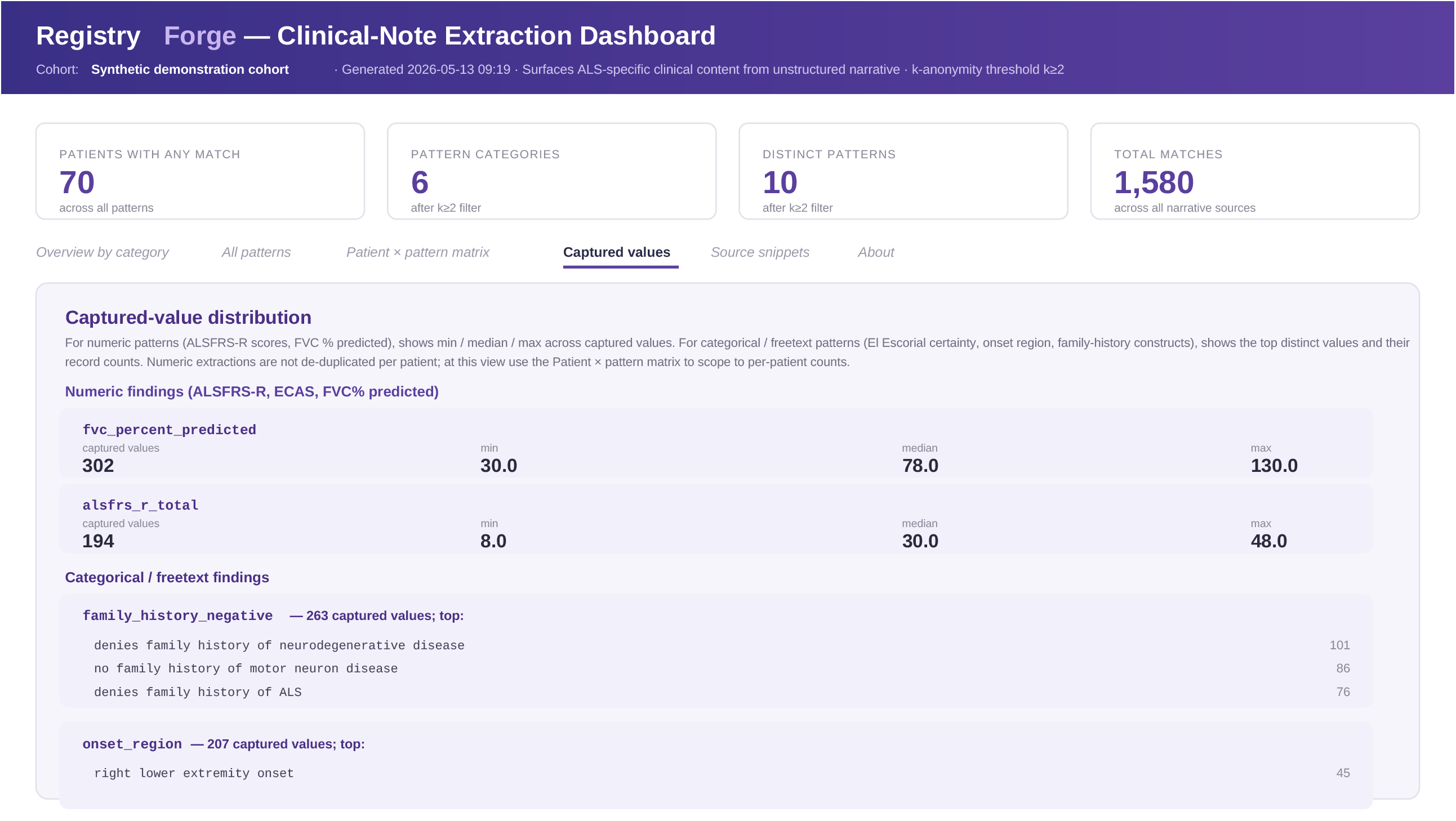

